# Parental educational attainment polygenic scores contribute to phenotypic heterogeneity in offspring with autism

**DOI:** 10.64898/2026.06.03.26354779

**Authors:** Shilin Gao, Yang Sui, Panhui Tian, Xiaoli Rao, Chenghao Yan, Yue Xu, Tianyun Wang

## Abstract

Educational attainment-related polygenic scores have been implicated in autism spectrum disorder (ASD), but how parental polygenic scores shape offspring phenotypes remains unclear. Using genotyping and exome-sequencing data from 142,357 individuals (55,252 ASD cases) in a large ASD cohort, we dissected the direct and indirect genetic effects of educational attainment-related polygenic scores on ASD phenotypes. Trio-model analyses showed that parental polygenic scores for educational attainment (PGS_EA_) were associated with milder core ASD symptoms, including social deficits and repetitive behaviors, predominantly through indirect genetic effects, whereas their associations with comorbidities were driven predominantly by direct genetic effects. PGS_EA_ was also significantly negatively associated with rare variant burden and prenatal factors, although these factors contributed largely independently to most phenotypes. Adjustment for full-scale intelligence quotient (FSIQ) and socioeconomic status (SES) partially attenuated the indirect effects of PGS_EA_ on offspring phenotypes. Finally, higher parental PGS_EA_ was associated with later age at diagnosis in offspring, partly through its protective effects on ASD phenotypes. These findings indicate that indirect genetic effects of parental PGS_EA_ contribute substantially to phenotypic variation in ASD and highlight family-mediated pathways as an important component of ASD heterogeneity.

## Introduction

Autism spectrum disorder (ASD) is a highly heterogeneous neurodevelopmental condition^1^. In addition to the core features of social communication deficits and restricted, repetitive behaviors, individuals with ASD frequently present with a range of comorbidities, including intellectual disability, sleep disorders and schizophrenia^2^. Consistent with this clinical heterogeneity, the genetic architecture of ASD is also highly complex^3^. In addition to rare *de novo* single-nucleotide variants/indels^4–7^, copy number variants^8–10^ and rare inherited loss-of-function variants^11–14^, common variants^15^ also contribute substantially to disease risk and phenotypic variation. Polygenic studies have shown that polygenic scores for ASD, schizophrenia and educational attainment (PGS_EA_) are all significantly associated with ASD^16^. Among these, PGS_EA_ has attracted particular attention because of their distinctive relationship with ASD phenotypes^17^. Previous studies have shown that PGS_EA_ is overtransmitted to individuals with ASD and is associated with milder clinical presentation^17,18^. However, existing work has focused primarily on the association between proband PGS_EA_ and ASD phenotypes, whereas the potential contribution of parental PGS_EA_ to offspring phenotypic variation remains unclear. PGS_EA_ can also be decomposed into a cognitive component, represented by the polygenic score for cognitive performance (PGS_CP_), and a non-cognitive component, represented by the polygenic score for non-cognitive skills (PGS_NON_COG_), which captures traits such as motivation, curiosity, persistence and self-control^19^. However, their associations with ASD are still unknown.

The concept of “genetic nurture” has been proposed to describe the phenomenon whereby non-transmitted parental alleles can influence offspring phenotypes indirectly by shaping the family environment or other early-life exposures, including prenatal factors^20^. In contrast, the effects of transmitted alleles on offspring phenotypes are referred to as direct genetic effects. Indirect genetic effects of PGS_EA_ on educational attainment and cognitive ability have been demonstrated in previous studies^20,21^. In addition, recent work in developmental disorders (DD) has suggested that PGS_EA_ may influence disease risk through parental non-transmitted effects, potentially reflecting indirect genetic effects^22^. Notably, although DD and ASD share a substantial rare variant burden, the contribution of common variation appears to differ between the two conditions. For example, PGS_EA_ is undertransmitted in DD^22^ but overtransmitted in ASD^16^, suggesting that educational attainment-related polygenic liability may have distinct implications across neurodevelopmental disorders. These observations highlight the need to determine whether PGS_EA_ influences phenotypic variation in ASD primarily through direct or indirect genetic effects, and through which pathways. In addition to genetic factors, environmental exposures such as prematurity have also been associated with both disease risk and phenotypic variation in neurodevelopmental disorders^23,24^. However, how these exposures interact with PGS_EA_ in shaping ASD phenotypes remains unclear.

In this study, we used genotyping and exome-sequencing data from 142,357 individuals (55,252 ASD cases) in the Simons Foundation Powering Autism Research for Knowledge (SPARK) cohort^25^ to investigate the association between PGS_EA_ and ASD phenotypes from the perspective of genetic nurture. These findings provide new insights into the phenotypic heterogeneity of ASD.

## Results

### Educational attainment-related polygenic scores are associated with milder ASD phenotypes

We constructed three polygenic scores related to educational attainment: PGS_EA_ (educational attainment), PGS_CP_ (cognitive performance) and PGS_NON_COG_ (the non-cognitive component of educational attainment). Polygenic transmission disequilibrium testing showed significant overtransmission of all three scores to ASD probands (Fig. S1, Supplementary Table 1), whereas no such overtransmission was observed in their unaffected siblings, consistent with previous studies^16^. The transmission deviation followed a clear gradient, with PGS_EA_ showing the strongest overtransmission, followed by PGS_CP_ and then PGS_NON_COG_. This pattern remained after stratification by sex. Although PGS_CP_ reached only nominal significance in females (*P_adj_* = 0.069), this was likely due to the smaller sample size in this subgroup. Together, these findings further support an important contribution of educational attainment-related polygenic liability to ASD risk.

To further characterize the relationship between educational attainment-related polygenic scores and ASD phenotypes, we regressed phenotypic measures in ASD probands, including the Social Communication Questionnaire (SCQ), Social Responsiveness Scale (SRS), Repetitive Behavior Scale-Revised (RBSR), Vineland Adaptive Behavior Scales (VABS), Developmental Coordination Disorder Questionnaire (DCDQ), FSIQ and comorbidities. In this population model, each regression included a single polygenic score and was adjusted for sex, age and the first 10 principal components. In probands, higher PGS_EA_ was associated with higher FSIQ, DCDQ and VABS scores, as well as lower SCQ, SRS and RBSR scores, indicating a broadly milder phenotypic profile (Fig. 1a, Supplementary Table 2). Similar patterns were observed for PGS_CP_ and PGS_NON_COG_, although the effect sizes were generally smaller than those for PGS_EA_. Higher PGS_EA_ in probands was also consistently associated with a lower risk of ASD comorbidities (Developmental disorders, OR: 0.89-0.94, *P_adj_* =7.98 × 10^−12^; Mood disorders, OR: 0.86-0.90, *P_adj_* = 4.46 × 10^−20^; Behavioral disorders, OR: 0.80-0.84, *P_adj_* = 1.23 × 10^−49^; Schizophrenia, OR: 0.65-0.79, *P_adj_* = 1.09 × 10^−10^; Sleep disorders, OR: 0.82-0.88, *P_adj_* = 7.67 × 10^−16^; Fig. 1b, Supplementary Table 3). Together, these results indicate that higher educational attainment-related polygenic scores were associated with milder ASD phenotypes and lower comorbidity risk across multiple domains.

**Figure 1.**
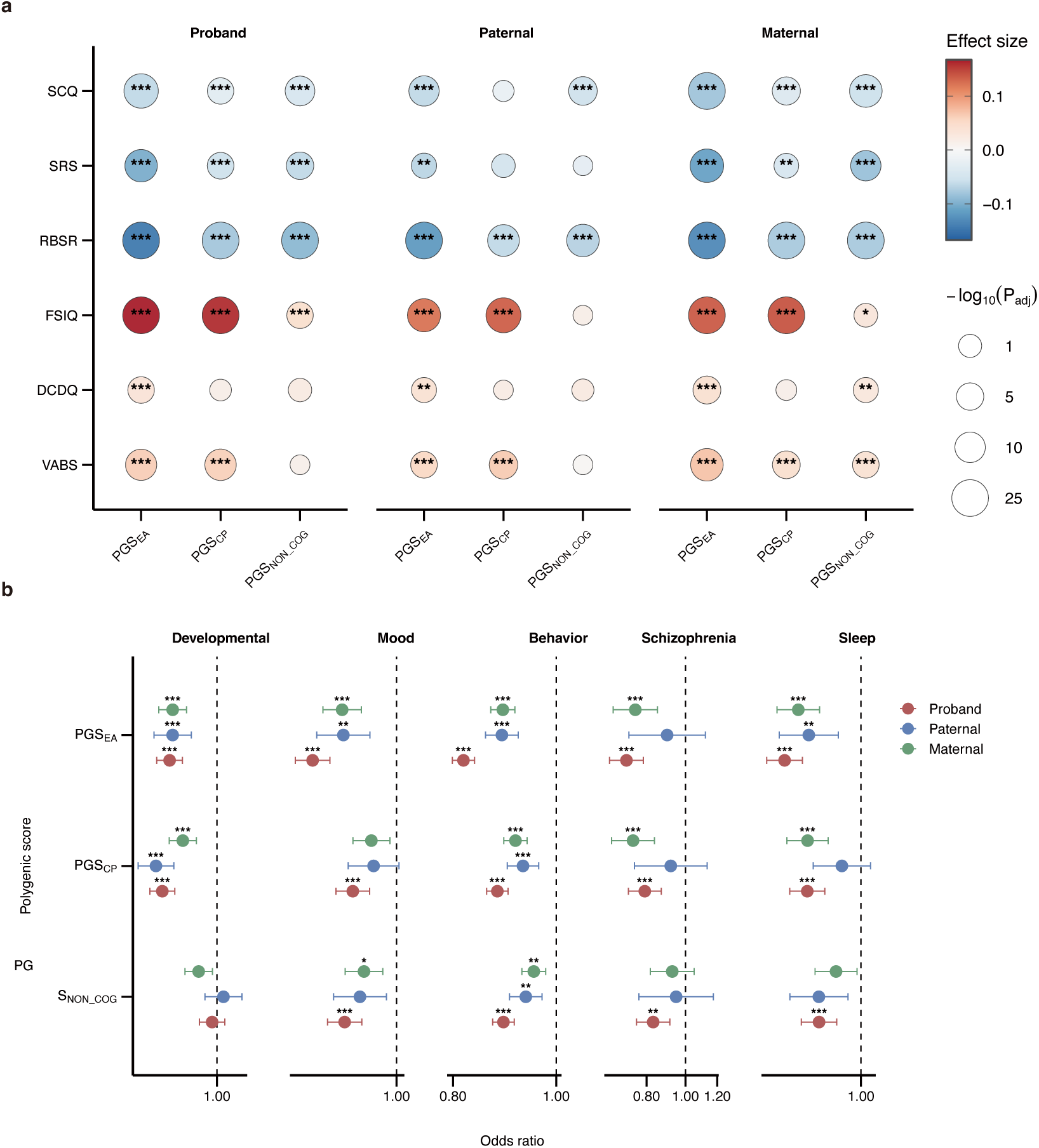
Associations of educational attainment-related polygenic scores with ASD phenotypes and comorbidities in the population model of full cohort. **a,** Associations of proband, paternal and maternal PGS_EA_, PGS_CP_ and PGS_NON_COG_ with quantitative ASD phenotypes in affected offspring in the population model. Effect estimates were obtained from linear regression models including a single polygenic score and meta-analyzed across the discovery and replication cohorts. Color indicates effect size and point size indicates −*log*_10_(*P_adj_*). b, Associations of proband, paternal and maternal PGS_EA_, PGS_CP_ and PGS_NON_COG_ with ASD comorbidities in affected offspring in the population model. Points indicate odds ratios and error bars indicate 95% confidence intervals. Asterisks denote Bonferroni-corrected significance levels: *P_adj_* < 0.05 (*); *P_adj_* < 0.01 (**); *P_adj_* < 0.001 (***).

Parental educational attainment-related polygenic scores showed similar associations with offspring phenotypes. Higher parental PGS_EA_ was associated with milder phenotypes in affected offspring (Fig. 1a, Supplementary Table 2; Fig. 1b, Supplementary Table 3). Notably, maternal PGS_NON_COG_ was significantly associated with higher offspring DCDQ and VABS scores, whereas no corresponding association was observed for offspring PGS_NON_COG_ itself. More broadly, associations between maternal polygenic scores and milder offspring phenotypes tended to be stronger than those observed for paternal scores. For example, higher maternal PGS_CP_ was significantly associated with lower SCQ (*P_adj_* = 9.85 × 10^−7^) and SRS (*P_adj_* = 8.34 × 10^−3^) scores in offspring, whereas comparable associations were not observed for paternal PGS_CP_. Analyses in trio samples yielded largely similar results (Fig. S2, Supplementary Table 4 and 5). Although some associations were attenuated and no longer reached significance, this was likely due to the smaller sample size. Overall, these findings suggest that educational attainment-related polygenic scores in both probands and parents were consistently associated with milder ASD phenotypes in affected offspring.

### Exploring the direct and indirect genetic effects

Although the population model identified robust associations between educational attainment-related polygenic scores and ASD phenotypes, it could not distinguish direct genetic effects in the child from indirect genetic effects mediated through parental genotypes and the family environment. To address this, we fitted trio models that simultaneously included polygenic scores for the child, father and mother. In this framework, the coefficient for the child polygenic score captures the direct genetic effect, whereas the coefficients for the parental polygenic scores capture indirect genetic effects acting through the family environment. PGS_EA_ showed significant protective associations with SCQ, SRS, RBSR and VABS, and these associations were driven predominantly by indirect rather than direct genetic effects (Fig. 2a, Supplementary Table 6). PGS_CP_ and PGS_NON_COG_ showed similar patterns. For FSIQ, both PGS_EA_ and PGS_CP_ showed evidence of direct and indirect genetic effects, consistent with the strong correlation between these two scores, whereas no significant association was observed for PGS_NON_COG_ (Fig. 2a, Supplementary Table 6). Across most phenotypes, indirect effects were slightly stronger for maternal than paternal polygenic scores, although these differences were not statistically significant. By contrast, the association between PGS_EA_ and behavioral disorders was driven mainly by direct genetic effects (OR: 0.79-0.90, *P_adj_* = 5.78 × 10^−5^, Fig. 2b, Supplementary Table 7). And the direct genetic effects of PGS_EA_ on developmental disorders (OR: 0.87-0.99, *P* = 0.021), mood disorders (OR: 0.84-0.97, *P* = 0.006) and schizophrenia (OR: 0.39-0.92, *P* = 0.019) were also nominally significant. Nominally significant direct genetic effects were also observed for PGS_CP_ on developmental disorders (OR: 0.88-0.99, *P* = 0.029) and behavioral disorders (OR: 0.87-0.99, *P* = 0.019), and for PGS_NON_COG_ on mood disorders (OR: 0.86-0.99, *P* = 0.019) and behavioral disorders (OR: 0.84-0.95, *P* = 4.18 × 10^−4^).

**Figure 2.**
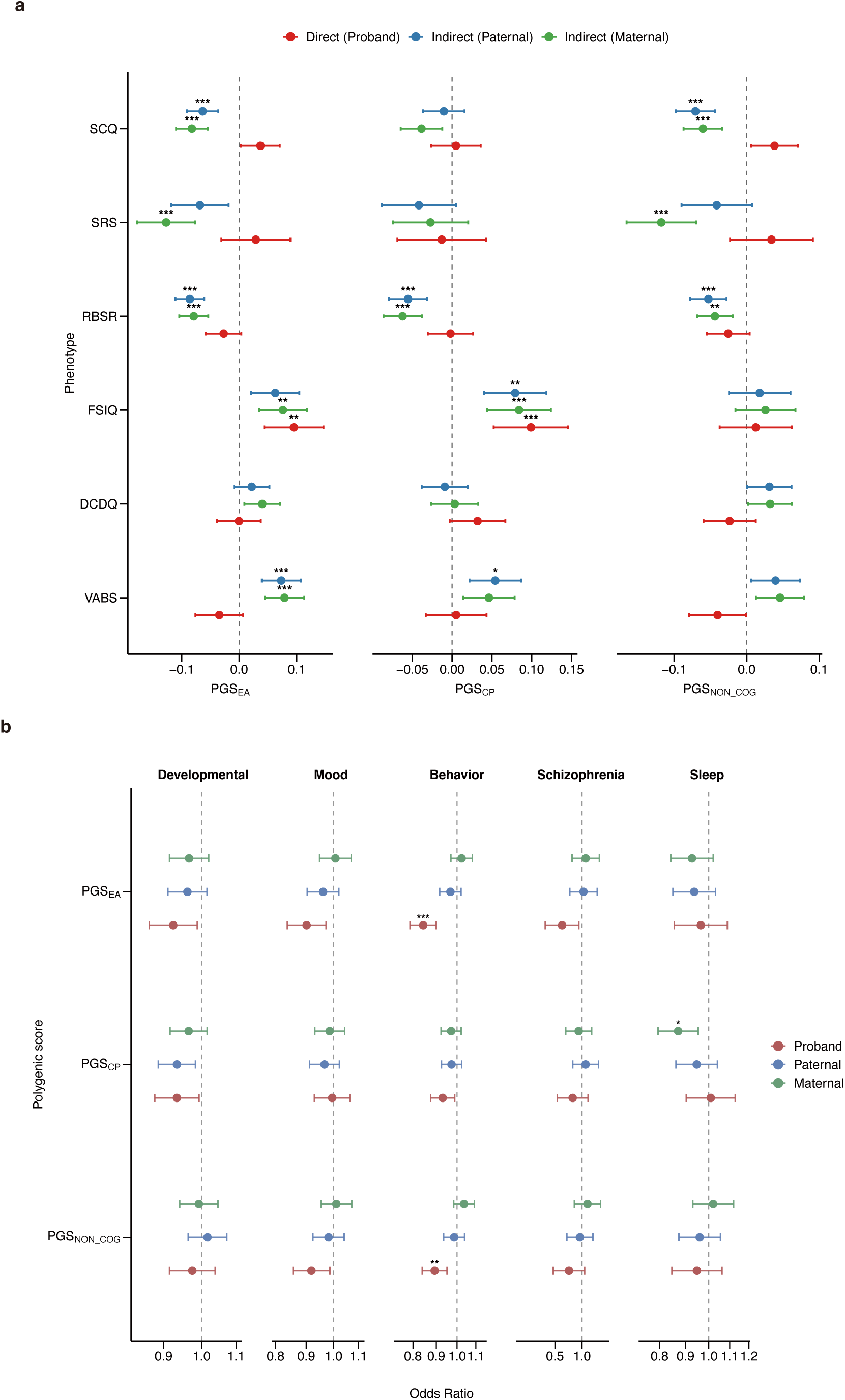
Direct and indirect genetic effects of educational attainment-related polygenic scores on ASD phenotypes and comorbidities in the trio model. **(a)** Associations of PGS_EA_, PGS_CP_, and PGS_NON_COG_ with ASD phenotypes in affected offspring in the trio model. The child coefficient represents the direct genetic effect, whereas the paternal and maternal coefficients represent indirect genetic effects. Effect estimates were obtained from linear regression models. Points indicate effect estimates and horizontal bars indicate 95% confidence intervals. **(b)** Associations of PGS_EA_, PGS_CP_, and PGS_NON_COG_ with comorbidities in the trio model. Effect estimates were obtained from logistic regression models. Points indicate odds ratios and error bars indicate 95% confidence intervals. Asterisks indicate Bonferroni-corrected significance thresholds: *P_adj_* < 0.05 (*); *P_adj_* < 0.01 (**); *P_adj_* < 0.001 (***).

These findings suggest that educational attainment-related polygenic scores influence core ASD phenotypes, including social and repetitive behaviors, primarily through indirect genetic effects, whereas their associations with comorbidities are more likely to arise through direct genetic effects.

### Interactions of rare variants and prenatal factors with direct and indirect genetic effects

In addition to common variants, rare variants have also been implicated in ASD phenotypic variation^17^. Previous studies have suggested that, under assortative mating^22^, rare and common variants may be correlated, potentially affecting the estimation of direct and indirect effects in the trio model. We therefore examined the relationship between rare variant burden and educational attainment-related polygenic scores, and tested whether adjustment for rare variants altered the estimated direct and indirect effects. PGS_EA_ was significantly negatively correlated with the damaging rare variant burden score within probands (r = −0.046, *P_adj_* = 1.90 × 10^−17^), within parents (r = −0.028, *P_adj_* = 1.02 × 10^−8^), and cross-parents (r = −0.025, *P_adj_* = 2.31 × 10^−4^) (Fig. 3a, Supplementary Table 8). PGS_NON_COG_ showed the same pattern and was significant in all three groups, whereas PGS_CP_ was significant only within probands. By contrast, no significant association was observed for the burden score based on synonymous variants.

**Figure 3.**
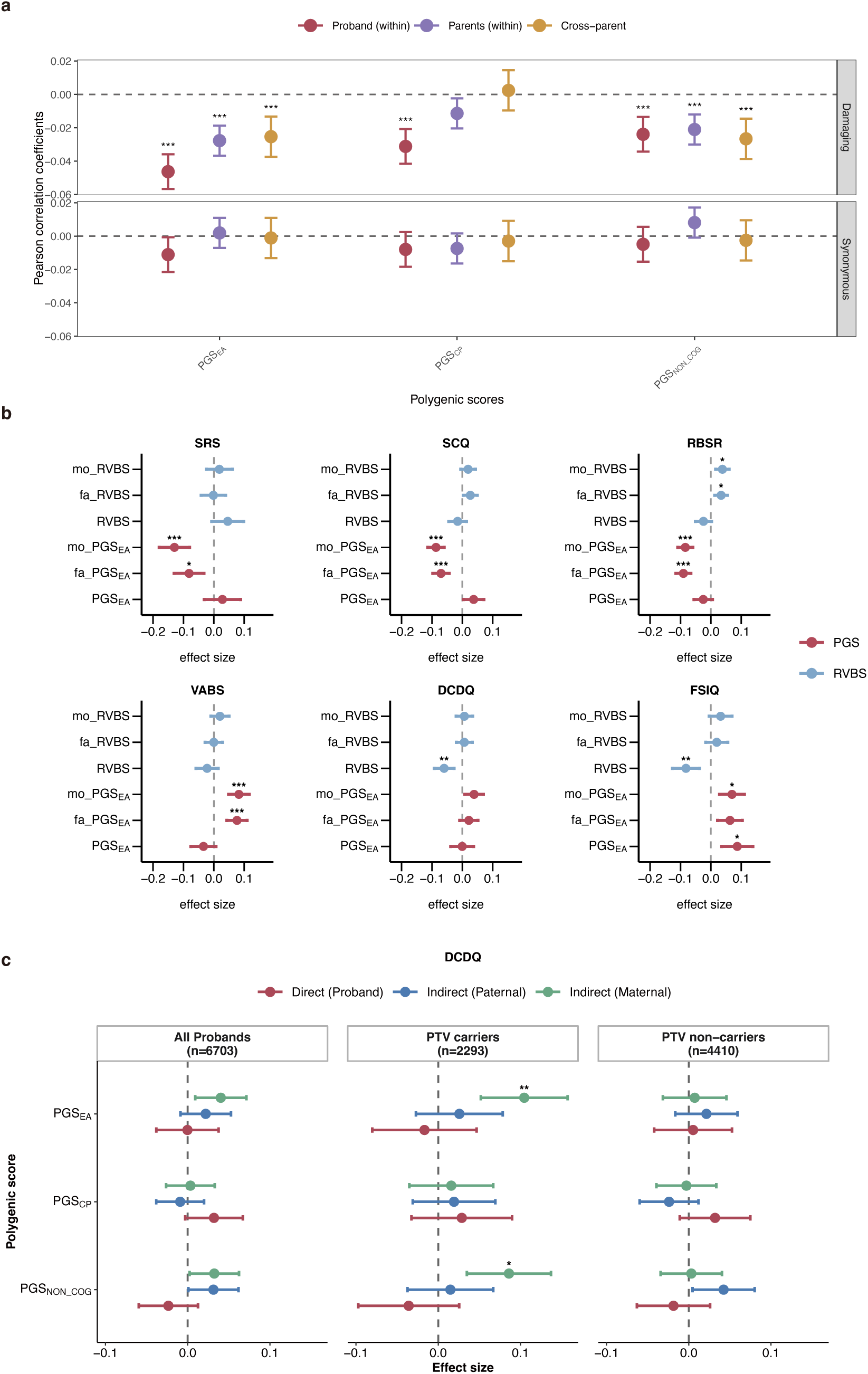
Association between rare variant burden and the effect of PGS in ASD. **a,** Correlations between polygenic scores and rare variant burden (RVBS). Points indicate Pearson correlation coefficients and error bars indicate 95% confidence intervals. **b,** The effects of PGS_EA_ in trio model after adjustment for rare variant burden. Effect estimates were obtained from linear regression models and meta-analyzed across the discovery and replication cohorts. Points indicate effect estimates and error bars indicate 95% confidence intervals. **c,** The effects of PGS on DCDQ in trio model stratified by PTV carrier status. Results are shown for all probands, PTV carriers, and PTV non-carriers. Points indicate effect estimates and error bars indicate 95% confidence intervals. Asterisks indicate Bonferroni-corrected significance thresholds: *P_adj_* < 0.05 (*); *P_adj_* < 0.01 (**); *P_adj_* < 0.001 (***).

We next asked whether the correlation between rare and common variants influenced the estimation of direct and indirect genetic effects. To account for rare variant burden, we extended the trio model by additionally including rare variant burden scores for the child, father and mother. Adjustment for rare variant burden did not materially alter the estimated direct or indirect effects of PGS_EA_ on ASD phenotypes and comorbidities (Fig. 3b, Fig. S3-5, Supplementary Table 9-10). As a complementary analysis, ASD probands were stratified by the presence or absence of protein-truncating variants (PTV) in SFARI genes^26^. The effects of PGS_EA_, PGS_CP_ and PGS_NON_COG_ on SRS, SCQ, RBSR, FSIQ and VABS were comparable between carriers and non-carriers (Fig. S6, Supplementary Table 11). For DCDQ, however, the indirect genetic effect of maternal PGS_EA_ was significantly associated with the phenotype in PTV carriers (n = 2,293, *P_adj_* = 0.0018) but not in non-carriers (n = 4,410, *P_adj_* = 1.00), despite the larger sample size of the non-carrier group (Fig. 3c, Supplementary Table 11). A similar pattern was observed for maternal PGS_NON_COG_. In line with this, the interaction term between maternal PGS_EA_ and PTV carrier status was nominally significantly associated with DCDQ (*P* = 0.0063, Supplementary Table 13), whereas no significant interaction was observed for the other phenotypes (Supplementary Table 13-14). These findings suggest a possible interaction between rare damaging variants and maternal educational attainment-related polygenic scores in motor coordination. Recent studies have implicated prenatal factors in variation in phenotypic severity^24^. PGS_EA_ has also been associated with lower prenatal risk in individuals with developmental delay^22^. We therefore examined whether prenatal factors modify the effects of educational attainment-related polygenic scores on ASD phenotypes. We considered several prenatal exposures in ASD probands, including fetal alcohol syndrome, bleeding into the brain, insufficient oxygen at birth, serious prenatal infection and premature birth. Higher PGS_EA_ and PGS_CP_ in probands were associated with lower prenatal risk (PGS_EA_, OR: 0.93-0.98, *P_adj_* = 0.0023; PGS_CP_, OR: 0.93-0.98, *P_adj_* = 0.0012; Fig. S7, Supplementary Table 15), whereas no significant association was observed for PGS_NON_COG_. However, adjustment for prenatal factors, or exclusion of individuals with prenatal factors, did not materially change the estimated effects of PGS_EA_ on ASD phenotypes, and similar results were observed for PGS_CP_ and PGS_NON_COG_ (Fig. S8, Supplementary Table 16). Consistent with this, no significant interaction between prenatal factors and polygenic scores was identified (Supplementary Table 17). These results suggest that prenatal factors and educational attainment-related polygenic scores exert independent effects on ASD phenotypes.

### FSIQ and household income partly account for parental PGS effects on ASD phenotypes

We next asked why parental educational attainment-related polygenic scores were associated with milder ASD phenotypes in offspring. Given the direct relationship between PGS_EA_ and FSIQ, we hypothesized that part of the protective effect of parental polygenic scores might be mediated through offspring FSIQ. Consistent with previous reports^18^, higher FSIQ was associated with lower SCQ, SRS and RBSR scores and with higher VABS and DCDQ scores in our data (Fig. 4a). We therefore compared parental PGS_EA_ effects on offspring phenotypes before and after adjustment for FSIQ. After adjustment, the associations of maternal PGS_EA_ with VABS and of paternal PGS_EA_ with SRS and SCQ were no longer significant (Fig. 4b, Supplementary Table 18). In line with this, the effects of parental polygenic scores on offspring VABS were positively correlated with their effects on offspring FSIQ (Pearson r =0.865, *P* = 0.0026; Fig. 4c). Together, these findings suggest that FSIQ partly accounts for the protective associations between parental polygenic scores and offspring ASD phenotypes.

**Figure 4.**
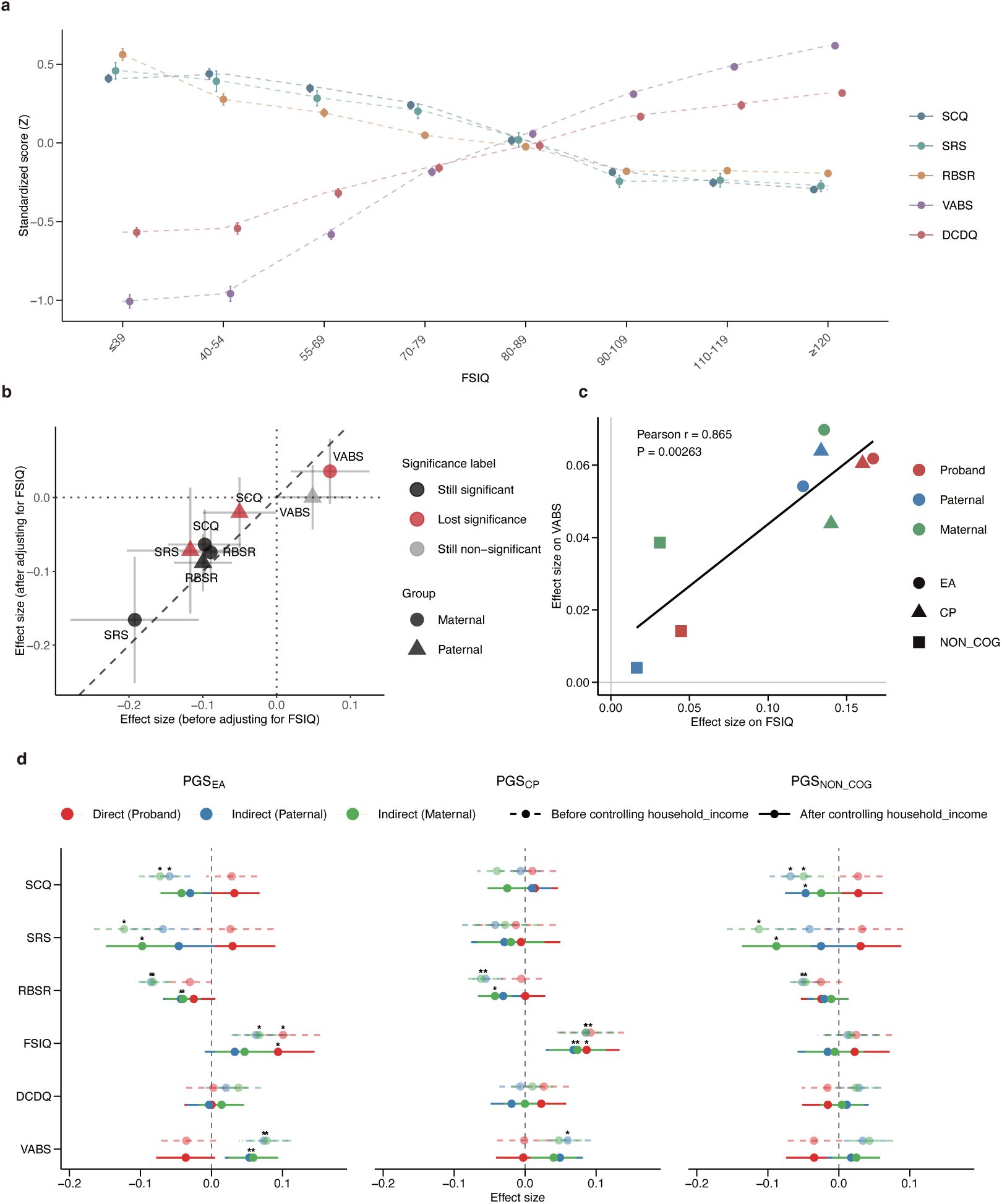
FSIQ and household income partly account for associations between parental polygenic scores and offspring ASD phenotypes. **a,** Standardized ASD phenotypic scores across FSIQ bins in ASD probands. **b,** Associations of parental PGS_EA_ with offspring phenotypes before and after adjustment for FSIQ. Points indicate effect estimates and horizontal and vertical error bars indicate 95% confidence intervals before and after adjustment, respectively. Point color denotes whether the association remained significant, lost significance, or remained non-significant after adjustment, and point shape indicates maternal or paternal effects. **c,** Correlation between the effects of parental polygenic scores on offspring FSIQ and their effects on offspring VABS in trio model.The solid line indicates the fitted linear relationship. **d,** Associations of PGS_EA_, PGS_CP_, and PGS_NON_COG_ with ASD phenotypes before and after adjustment for household income in the trio model. Asterisks indicate Bonferroni-corrected significance thresholds: *P_adj_* < 0.05 (*); *P_adj_* < 0.01 (**); *P_adj_* < 0.001 (***).

Socioeconomic status is closely related to educational attainment and has also been linked to ASD phenotypes such as motor coordination ^27^. We next examined whether socioeconomic status modifies the associations between educational attainment-related polygenic scores and ASD phenotypes. Higher parental PGS_EA_, PGS_CP_ and PGS_NON_COG_ were all significantly associated with higher household income (Supplementary Table 19). Adjustment for household income attenuated several indirect effects of parental polygenic scores on offspring phenotypes, some of which were no longer significant (Fig. 4d, Supplementary Table 20). The largest attenuation was observed for the indirect effect of PGS_NON_COG_ on RBSR (Maternal PGS_NON_COG_, before controlling: effect size = −0.047, *P_adj_* = 0.004; after controlling: effect size = −0.011, *P_adj_* = 1.00. Paternal PGS_NON_COG_, before controlling: effect size = −0.051, *P_adj_* = 6.97 × 10^−5^; after controlling: effect size = −0.021, *P_adj_* = 1.00). These results suggest that household income may partially account for the influence of parental polygenic scores on offspring phenotype. We further tested whether parental polygenic scores were associated with increased intervention in offspring. However, no significant association was observed between parental polygenic scores and offspring intervention status (Supplementary Table 21).

### Association between age at diagnosis and parental PGS_EA_

Recent studies have suggested that ASD cases diagnosed at different ages may differ in their polygenic architecture^28^. We therefore examined whether educational attainment-related polygenic scores were associated with age at diagnosis. Trio-model analyses showed that higher PGS_EA_ was significantly associated with later diagnosis, predominantly through indirect rather than direct genetic effects (Fig. 5a, Supplementary Table 22). Given the protective associations of PGS_EA_ with multiple ASD phenotypes, we hypothesized that higher PGS_EA_ might delay diagnosis by attenuating symptom severity. Mediation analyses supported this hypothesis: both paternal and maternal PGS_EA_ showed significant indirect effects on age at diagnosis through SCQ, SRS, RBSR, FSIQ and VABS (Fig. 5b, Supplementary Table 23). Notably, both maternal and paternal PGS_EA_ and PGS_CP_ showed significant mediated effects through offspring FSIQ, whereas only the unmediated effect of paternal PGS_CP_ reached nominal significance. Together, these findings suggest that educational attainment-related polygenic scores may delay ASD diagnosis in part by contributing to a milder phenotypic profile.

**Figure 5.**
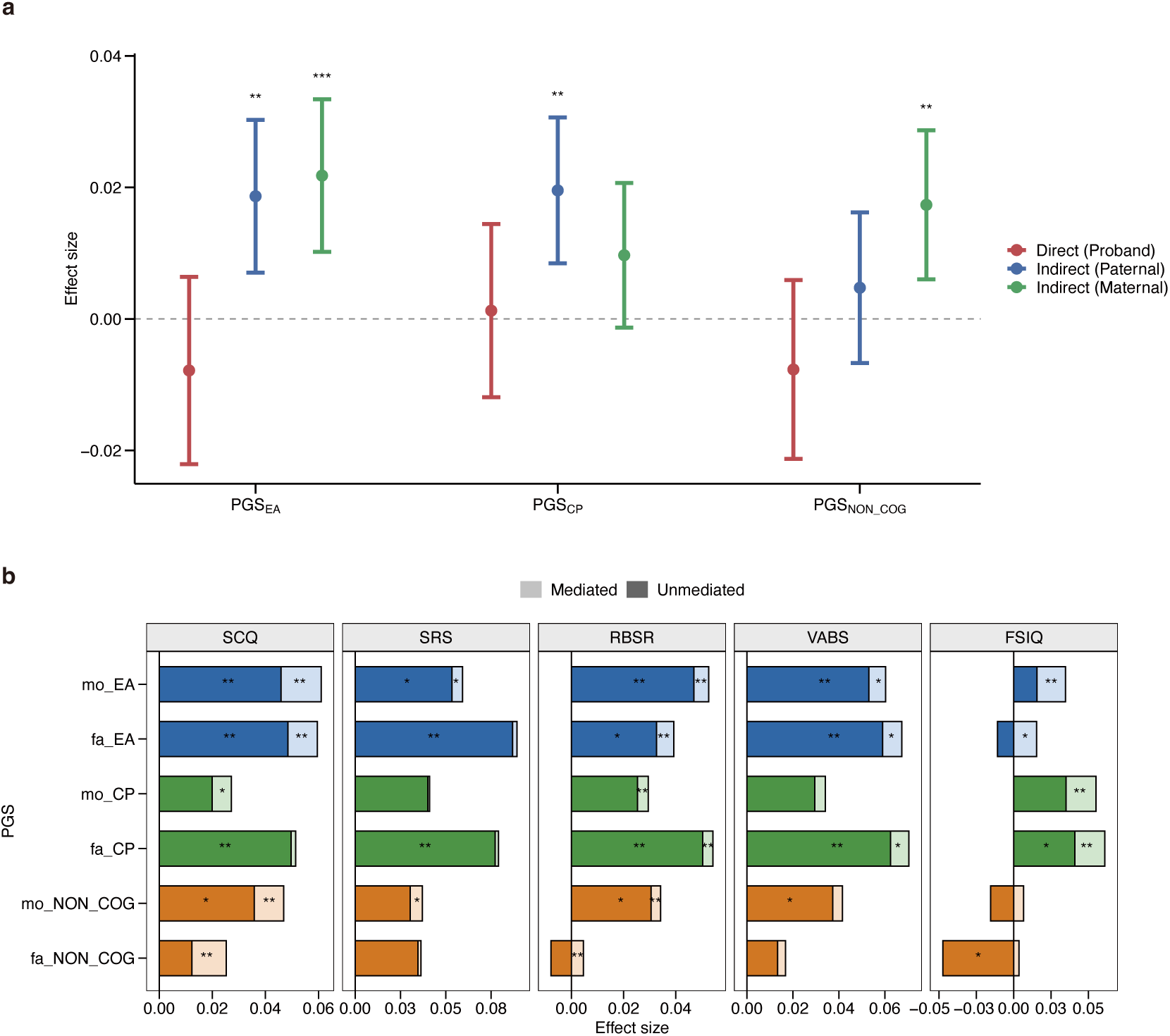
Educational attainment-related polygenic scores are associated with age at diagnosis and mediated phenotypic pathways. **a,** Associations of of PGS_EA_, PGS_CP_, and PGS_NON_COG_ with age at ASD diagnosis in the trio model. Points indicate effect estimates and vertical bars indicate 95% confidence intervals. **b,** Mediation analyses of the associations between parental educational attainment-related polygenic scores and age at diagnosis through SCQ, SRS, RBSR, VABS and FSIQ. Dark bars indicate unmediated effects and light bars indicate mediated effects. Effect estimates were meta-analyzed across the discovery and replication cohorts. Asterisks indicate significance: *P* < 0.05 (*); *P_adj_* < 0.05 (**);

Recent studies have suggested that the sex ratio approaches 1:1 among ASD cases diagnosed later in development^29^, a pattern that was also evident in our data (Fig. S9a, Fig. S9b). We therefore tested whether the effects of educational attainment-related polygenic scores on offspring phenotypes differed by sex. Trio-model analyses performed separately in males and females yielded broadly similar effect estimates, and no sex interaction term was significant (Fig. S9c, Supplementary Table 24-25). These findings suggest that the effects of educational attainment-related polygenic scores on ASD phenotypes are largely comparable between males and females.

## Discussion

Although educational attainment-related polygenic scores are known to be associated with ASD, how they contribute to phenotypic variation has remained unclear. Using genotyping data from a large ASD cohort, we systematically dissected their direct and indirect effects on ASD phenotypes.

Polygenic transmission disequilibrium testing confirmed that PGS_EA_ is overtransmitted to ASD probands, in line with previous studies^16^. We further found that both PGS_CP_ and PGS_NON_COG_ were also overtransmitted, suggesting that the association between educational attainment-related polygenic liability and ASD extends beyond cognitive components to include non-cognitive components (motivation, curiosity, persistence and self-control). Population-model analyses showed that higher PGS_EA_ in ASD probands was associated with milder ASD phenotypes and lower risk of comorbidities, and similar but weaker patterns were observed for parental polygenic scores. Compared with paternal scores, maternal polygenic scores tended to show broader associations with offspring phenotypes and larger effect sizes. Extending these observations, trio-model analyses suggested that the association of PGS_EA_ with core ASD phenotypes, including social deficits and repetitive behaviors, was driven mainly by indirect rather than direct genetic effects. In contrast, PGS_EA_ contributed to FSIQ through both direct and indirect pathways, whereas its associations with ASD comorbidities were largely direct. This pattern suggests substantial heterogeneity in the ways educational attainment-related polygenic liability relates to different dimensions of ASD. It also suggests that core ASD symptoms may be more sensitive than comorbidities to family- or environment-mediated influences, in line with previous evidence that family-based interventions can improve ASD symptoms^30,31^.

The indirect effects of PGS_EA_ on ASD phenotypes may be partly confounded by parental assortment. In the present study, educational attainment-related polygenic scores were negatively correlated with the burden of rare damaging variants. However, after adjustment for rare variant burden in the trio model, the estimated effects of PGS_EA_ remained largely stable across most phenotypes, suggesting that PGS_EA_ and rare damaging variation make largely independent contributions to ASD phenotypes. An exception to this overall pattern was observed for DCDQ, which captures developmental motor coordination. After adjustment for rare variant burden, the indirect effect of maternal PGS_EA_ on DCDQ reached nominal significance. Using a complementary approach, we further stratified ASD probands according to the presence or absence of PTVs in risk genes and found that the indirect effect of maternal PGS_EA_ on DCDQ was observed in PTV carriers but not in non-carriers. Consistent with this, the interaction between maternal PGS_EA_ and PTV carrier status was also nominally significant. Together, these findings suggest a potential interaction between rare damaging variants and indirect genetic effects in shaping motor function, a pattern that has not been reported in previous population-based analyses^16^.

Prenatal factors are also an important source of phenotypic heterogeneity in ASD. Previous studies have suggested that prematurity is associated with fewer ASD-associated mutations in ASD cases, and that its phenotypic effects are largely independent of those of rare variants^24^. In the present study, higher PGS_EA_ was associated with lower prenatal risk. However, neither adjustment for prenatal factors nor exclusion of individuals with prenatal exposures materially altered the estimated effects of PGS_EA_ in the trio model. These findings suggest that prenatal factors are unlikely to account for the indirect effects of PGS_EA_ on ASD phenotypes.

Consistent with both previous reports^18^ and our findings, other ASD-related phenotypes were attenuated with increasing FSIQ levels. Because parental PGS_EA_ showed relatively strong indirect genetic effects on offspring FSIQ, together with a significant direct genetic effect, we reasoned that its substantial influence on IQ might have a knock-on effect on other phenotypes, such as social deficits and repetitive behaviors. We therefore repeated the trio-model analyses with additional adjustment for FSIQ. After adjustment for FSIQ, several indirect effects of PGS_EA_ on offspring phenotypes were no longer significant. Consistent with this, the effects of parental polygenic scores on offspring VABS were positively correlated with their effects on offspring FSIQ. Together, these findings suggest that differences in FSIQ may partly account for the associations of PGS_EA_ with multiple offspring phenotypes. They also indicate that complex ASD phenotypes are closely interrelated, with variation in one domain often accompanied by variation in others.

Previous studies have shown that socioeconomic status is significantly associated with ASD phenotypes and is also strongly related to PGS_EA_^27,32,33^. We therefore explored the possibility that PGS_EA_ may improve offspring phenotypes in part through increasing socioeconomic status. After adjustment for household income, several indirect genetic effects of parental polygenic scores on offspring phenotypes were no longer significant. These findings suggest that socioeconomic status may represent one potential pathway through which parental polygenic scores exert indirect genetic effects.

Finally, we investigated the relationship between the indirect effects of parental PGS_EA_ and age at ASD diagnosis. Unexpectedly, higher parental PGS_EA_ was significantly associated with later diagnosis. Recent studies have likewise suggested that common genetic variation contributes substantially to variation in age at diagnosis^28^. We further found that the association between PGS_EA_ and later diagnosis was partly mediated through its effects on ASD phenotypes. These findings provide a new perspective for understanding variation in age at diagnosis among individuals with ASD.

Several limitations of the present study should be noted. First, all analyses were conducted in individuals diagnosed with ASD, and the present findings may not generalize to individuals in the general population who exhibit subclinical autistic traits but do not meet diagnostic criteria. Previous studies have suggested that the same genetic variants may have different phenotypic effects in affected individuals and in the general population^34^. Second, because the present study was based on cross-sectional rather than longitudinal data, we were unable to assess how the effects of PGS_EA_ on ASD phenotypes may change over development^21^. Nevertheless, our findings provide a new perspective on the phenotypic heterogeneity of ASD and highlight family-mediated pathways as potentially relevant to ASD-related phenotypes.

## Methods

### Datasets

We analyzed data from the SPARK cohort using iWES v3, released in August 2024. This dataset includes 142,357 individuals from 54,558 families, with exome sequencing data available for 142,357 individuals and genotyping data for 141,368 individuals. The cohort comprised 55,252 individuals with ASD and 87,105 family members, including siblings, fathers, mothers and other relatives. Phenotypic information was obtained from SPARK Collection V18, released in December 2025.

### Phenotypes

The phenotypes included in this study were the Social Communication Questionnaire (SCQ), Social Responsiveness Scale (SRS), Repetitive Behavior Scale-Revised (RBSR), Vineland Adaptive Behavior Scales (VABS), full-scale intelligence quotient (FSIQ) and Developmental Coordination Disorder Questionnaire (DCDQ). We also included ASD comorbidities, including developmental disorders, mood disorders, sleep disorders, behavioral disorders and schizophrenia. Developmental disorders included speech and language disorder, intellectual disability/cognitive impairment, learning disability, and other developmental delay or developmental disability. Mood disorders included mood disorder, depression, anxiety and obsessive-compulsive disorder. Behavioral disorders included attention-deficit/hyperactivity disorder, conduct disorder, intermittent explosive disorder and oppositional defiant disorder. These comorbidity data were obtained from the “Basic Medical Screening” module of the SPARK dataset and were analyzed as binary phenotypes.

### Genotype quality control and imputation

Genotype quality control, relatedness inference, principal-component analysis and imputation were performed broadly following the SPARK workflow described by Zhang et al^28^. Samples from SPARK WES batches 1–4 were genotyped on the Illumina Global Screening Array v2 and were used as the discovery cohort (n = 70,487), whereas samples from WES batches 5–9 were genotyped using genotyping-by-sequencing and were used as the replication cohort (n = 71,267). Analyses were restricted to individuals of European ancestry according to SPARK annotations. At the sample level, individuals were excluded if genotype call rate was below 98%, if reported sex was inconsistent with genetically inferred sex, or if heterozygosity was more than 3 standard deviations from the cohort mean. In families with trio data, trios with Mendelian error rates greater than 5% were removed. At the variant level, markers were retained if minor allele frequency exceeded 1%, call rate exceeded 95%, and Hardy–Weinberg equilibrium *P* values were greater than 1 × 10^−6^. Pairwise relatedness was estimated using KING^35^. For principal-component analysis, variants were pruned for linkage disequilibrium at r^2^ < 0.1, and the major histocompatibility complex region was excluded. Principal components were then computed in genetically unrelated individuals using PC-AiR^36^ implemented in GENESIS (v2.22.2) and projected onto related individuals. Imputation was performed on the TOPMed Imputation Server^37,38^ (v1.7.3) using the TOPMed reference panel, with Eagle v2.5 for phasing and Minimac4^38^ for imputation. After imputation, variants with minor allele frequency >0.1% and imputation quality R^2^ > 0.6 were retained for downstream analyses.

### Polygenic score construction

Polygenic scores were generated using PRS-CS^39^ (v1.1.0), which uses a Bayesian continuous shrinkage prior to infer posterior SNP effect sizes from GWAS summary statistics. Three educational attainment-related PGSs were constructed, including PGS_EA_, PGS_CP_ and PGS_NON_COG_. PGS_EA_ was based on the GWAS meta-analysis of educational attainment by Lee et al^40^, which included approximately 1.1 million individuals. PGS_CP_ and PGS_NON_COG_ were based on the study by Demange et al^19^, which used GWAS-by-subtraction to separate the cognitive and non-cognitive genetic components of educational attainment. PGSs were calculated from imputed genotype data and standardized prior to association analyses.

### Rare variant processing and annotation

Per-sample pVCF files generated by GATK in the SPARK dataset were downloaded for rare variant analyses. Variant processing was conducted in Hail^41^ v0.2 to generate the analysis-ready dataset. After import, multiallelic variants were decomposed into biallelic records, and low-complexity regions were excluded. Sample sex was inferred using the impute_sex() function in Hail, after which a series of genotype-level filters were applied. Genotype calls were removed if sequencing depth was <10 or >1000, except in male hemizygous regions, where calls with depth <7 were excluded. Additional filtering was performed based on genotype quality metrics. Homozygous reference calls were removed when genotype quality was <25, whereas heterozygous and homozygous alternate calls were removed when the phred-scaled likelihood for the homozygous reference genotype was <25. Heterozygous calls falling in hemizygous regions in males were excluded, and calls on chromosome Y in female samples were also removed. For heterozygous calls, genotypes were further filtered if allele balance was <0.25, if the binomial-test probability for the observed allele balance was <1 × 10^−9^, or if the number of informative reads supporting the genotype was <90% of total depth. A similar informative-read filter was applied to homozygous alternate calls. At the variant level, sites with call rate <10% or Hardy–Weinberg equilibrium *P* < 1 × 10^−12^ were excluded. Rare variants were defined as variants with allele frequency <0.001 in both our dataset and the gnomAD v2.1 exome, together with AC <5.

### Polygenic transmission disequilibrium tests

Polygenic transmission disequilibrium tests (pTDT) were performed to evaluate whether educational attainment-related polygenic scores were overtransmitted to ASD probands relative to expectation from parental genotypes. Analyses were carried out separately in the discovery and replication cohorts and subsequently combined by meta-analysis. Three polygenic scores were tested, including PGS_EA_, PGS_CP_ and PGS_NON_COG_. Within each family, the mid-parent polygenic score was calculated as the average of the paternal and maternal scores. For each offspring, the pTDT deviation was defined as the difference between the offspring polygenic score and the corresponding mid-parent value, divided by the standard deviation of the mid-parent score in the analyzed sample. For each polygenic score, the mean deviation was tested against zero using a one-sample t-test. Positive deviation indicates overtransmission, whereas negative deviation indicates undertransmission. Analyses were conducted separately in ASD probands and unaffected siblings. In addition to the overall sample, sex-stratified analyses were performed in males and females.

### Effects of educational attainment-related polygenic scores on ASD phenotypes

Associations between educational attainment-related polygenic scores and ASD phenotypes were evaluated in ASD probands of European ancestry from the discovery and replication cohorts separately, followed by meta-analysis across cohorts. Continuous outcomes, including SCQ, SRS, RBSR, VABS, FSIQ and DCDQ, were analyzed using linear regression, whereas binary outcomes, including comorbidities and other dichotomous phenotypes, were analyzed using logistic regression. Unless otherwise specified, continuous phenotypes and polygenic scores were standardized within cohort to mean 0 and standard deviation 1 before analysis.

For the population model, a single polygenic score was entered into the regression model at a time. Analyses of offspring polygenic scores included the offspring score as the predictor of interest and were adjusted for sex, phenotype-specific age when available, and the first 10 ancestry principal components of the offspring. Analyses of paternal and maternal polygenic scores were conducted analogously. The general form of the model for continuous phenotypes was

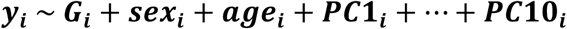

where ***y_i_*** denotes the phenotype and ***G_i_*** denotes the polygenic score under consideration. For binary outcomes, the same covariates were included in a logistic regression framework.

To decompose offspring and parental contributions, we fitted trio models in which the offspring, paternal and maternal polygenic scores for the same trait were entered jointly. In these models, the coefficient for the offspring polygenic score estimates the direct genetic effect, whereas the coefficients for the paternal and maternal polygenic scores capture non-transmitted parental effects, which may reflect indirect genetic effects as well as residual confounding. For continuous phenotypes, the trio model took the form

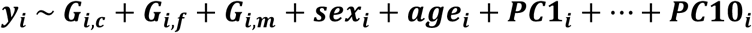

where *G_i,c_*, *G_i,f_* and *G_i,m_* denote the offspring, paternal and maternal polygenic scores, respectively. Logistic regression was used for dichotomous outcomes with the same covariate structure.

All association analyses were performed separately in the discovery and replication cohorts. Effect estimates and standard errors were then combined using meta-analysis.

### Analysis of rare coding variants

Variants were annotated using ANNOVAR. For gene-level analyses, one variant was retained per individual per gene, prioritizing the variant with the most severe predicted functional consequence. Frameshift variants, splice-acceptor variants, splice-donor variants and stop-gained variants were classified as protein-truncating variants (PTVs). Missense variants with MPC^42^ scores ≥2 were classified as MisB variants. PTVs and MisB variants were collectively defined as damaging variants. We then calculated a rare variant burden score as the number of damaging variants carried by each individual in constrained genes (pLI^43^ > 0.9). As a negative control, we additionally quantified the number of synonymous variants carried by each sample.

We tested whether rare variant burden was correlated with educational attainment-related polygenic scores by calculating Pearson correlation coefficients between rare variant burden scores and PGS_EA_, PGS_CP_ or PGS_NON_COG_. To account for ancestry, rare variant burden scores and polygenic scores were separately residualized on the first 10 genetic principal components before analysis. Correlations were evaluated within probands, within parents and across parents. In the within-parent analysis, paternal and maternal observations were combined and correlated with their own polygenic scores; in the cross-parent analysis, each parent’s rare variant burden was correlated with the partner’s polygenic score. Analyses were performed separately in the discovery and replication cohorts and combined by meta-analysis using Fisher’s z transformation.

To test whether rare damaging coding variation influenced the estimated effects of educational attainment-related polygenic scores on ASD phenotypes, we extended the trio model by additionally adjusting for RVBS n the child, father and mother. For each educational attainment-related polygenic score, the adjusted trio model included the offspring, paternal and maternal polygenic scores jointly, together with the corresponding child, paternal and maternal RVBS. The model for continuous phenotypes was:

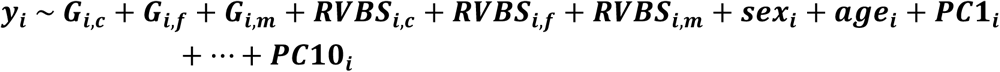

Where ***RVBS_i,c_***, ***RVBS_i,f_***, ***RVBS_i,m_*** denote the corresponding RVBS.

To assess whether the effects of educational attainment-related polygenic scores differed according to rare damaging variation in known ASD risk genes, we stratified ASD probands by the presence or absence of protein-truncating variants (PTVs) in SFARI genes. Trio models were then fitted separately in PTV carriers, non-carriers and the full ASD sample. To formally test whether trio-model effects differed by SFARI gene PTV status, we fitted interaction models in the full ASD sample

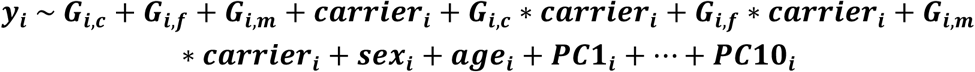

where ***carrier_i_*** denotes SFARI gene PTV carrier status for offspring i.

### Association of polygenic scores with prenatal factors

Associations between educational attainment-related polygenic scores and prenatal factors were evaluated in ASD probands using logistic regression. Prenatal factors were treated as binary outcomes. Offspring, paternal and maternal polygenic scores were tested separately in univariate models. Models were adjusted for sex, age and the first 10 principal components.

To test whether prenatal factors influenced the estimated effects of educational attainment-related polygenic scores on ASD phenotypes, we incorporated prenatal factors into trio-based association analyses using two complementary strategies. First, we repeated trio-model analyses after excluding exposed individuals for each prenatal factor separately, retaining only offspring with prenatal factor value equal to 0. Second, we repeated the same trio models while including each prenatal factor as an additional covariate. Prenatal factors used as covariates were coded strictly as binary variables (0/1). To further test whether prenatal factors modified polygenic-score effects, we fitted interaction models in the full ASD sample. For each prenatal factor, offspring, paternal and maternal polygenic scores were entered jointly together with the prenatal factor and the corresponding interaction terms.

### Analyses of FSIQ, household income and intervention status

To explore potential pathways linking parental educational attainment-related polygenic scores to offspring ASD phenotypes, we performed a series of follow-up analyses focusing on offspring FSIQ, household income and intervention status. We first assessed its association with ASD phenotypes using regression models with the same covariates as in the primary analyses. We then repeated the trio models after further adjustment for offspring FSIQ or household income and compared the parental polygenic-score effect estimates before and after this adjustment. To minimize differences driven by sample availability, these comparisons were additionally evaluated in matched subsets with complete data on FSIQ or household income.

### Mediation analyses of age at diagnosis

To test whether the association between parental educational attainment-related polygenic scores and age at diagnosis was mediated through ASD phenotypes, we performed mediation analyses using structural equation modeling. Separate models were fitted for each mediator, including SCQ, SRS, RBSR, VABS and FSIQ, and for each parental polygenic score. In each model, age at diagnosis was treated as the outcome, the selected phenotype as the mediator, and the parental polygenic score as the exposure of interest. The corresponding offspring and the other parental polygenic score were included as covariates, together with sex and the first 10 ancestry principal components. Continuous variables were standardized before analysis. The structural equation model was fit using the R package *laavan*. Models were fitted separately in the discovery and replication cohorts, and indirect, direct and total effects were subsequently combined by meta-analysis.

## Data Availability

The genomic and phenotypic data for the SPARK study (https://www.sfari.org/resource/spark/) are available at SFARI Base (https://base.sfari.org).

## Acknowledgements

We are grateful to all the participants and staff of SPARK for their invaluable contributions to this study. This work was supported by the Brain Science and Brain-like Intelligence Technology-National Science and Technology Major Project (2025ZD0218000), National Natural Science Foundation of China (82471194), and the “Fundamental Research Funds for the Central Universities” starting fund (BMU2022RCZX038) to T.W. We also thank Evan E. Eichler, Daniel S. Malawsky, Kaitlin E. Samocha, Dianzhi Liu, and Tian Ge for their helpful advice during the analyses and preparation of the manuscript.

## Competing interests

The authors declare no competing interests.

## Additional information

The genomic and phenotypic data for the SPARK study (https://www.sfari.org/resource/spark/) are available at SFARI Base (https://base.sfari.org) under request. The code relevant to quality control, imputation and genome-wide association analyses in SPARK is available at https://github.com/vwarrier/SPARK_iWES2_imputation/. Polygenic scores were generated using PRS-CS v1.1.0 which available at https://github.com/getian107/PRScs, and the variant processing was performed using Hail-based pipelines available at https://github.com/jsealock1/sequencing_qc.

## Supplementary Figures

**Figure S1.**
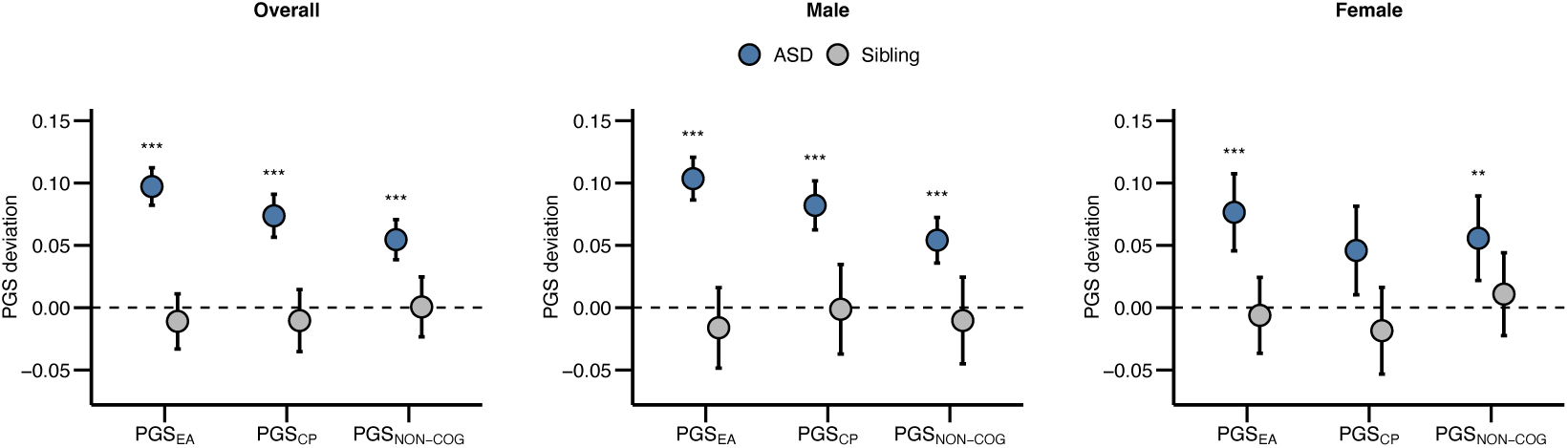
Overtransmission of educational attainment-related polygenic scores to ASD probands. Polygenic transmission disequilibrium test (pTDT) results for PGS_EA_, PGS_CP_ and PGS_NON_COG_ are shown for ASD probands and unaffected siblings in the overall sample and after stratification by sex. Estimates were meta-analyzed across the discovery and replication cohorts. Points indicate pTDT deviations and error bars indicate 95% confidence intervals. Asterisks denote Bonferroni-corrected significance levels: *P_adj_* < 0.05 (*); *P_adj_* < 0.01 (**); *P_adj_* < 0.001 (***).

**Figure S2.**
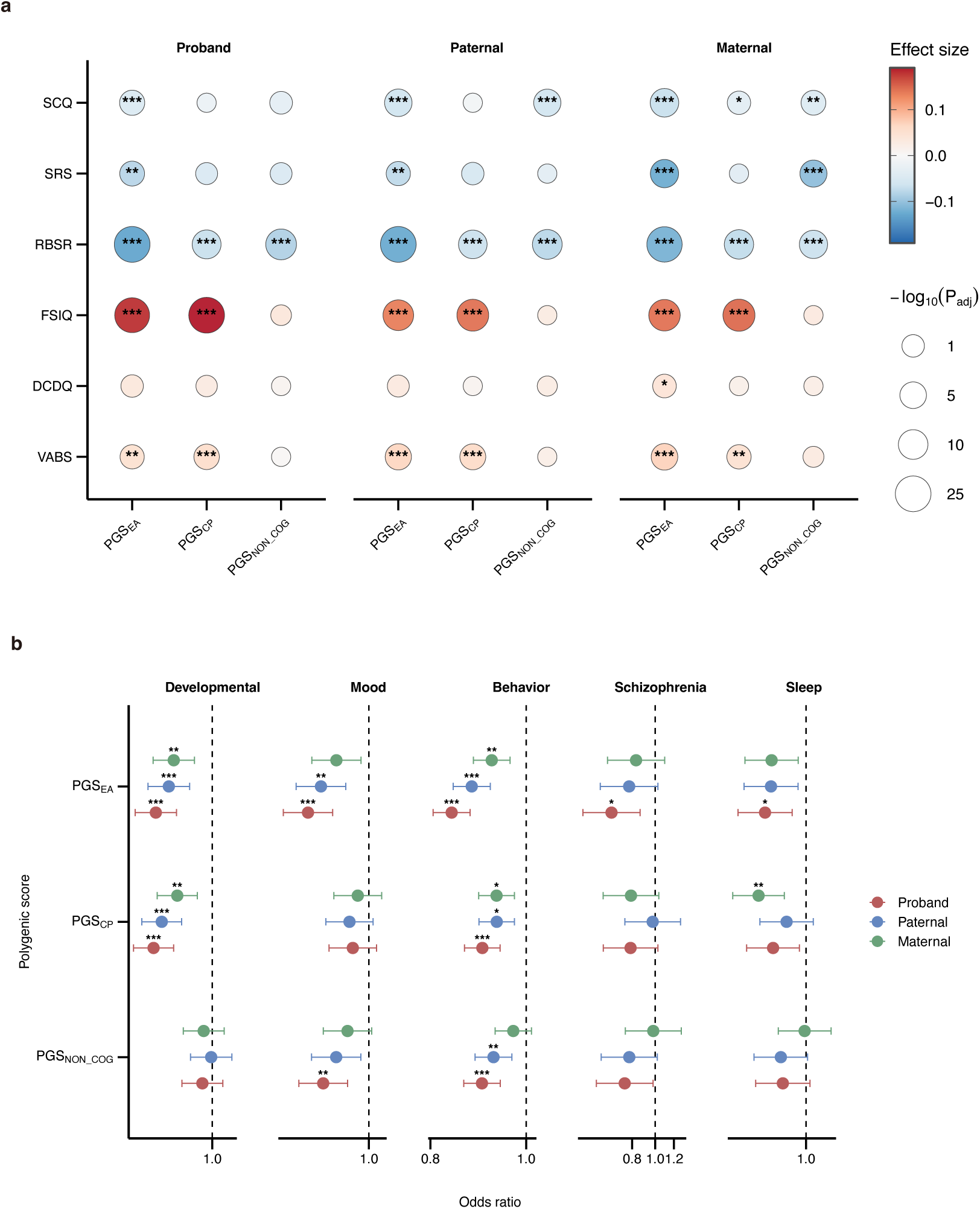
Associations of educational attainment-related polygenic scores with ASD phenotypes and comorbidities in the population model of trio sample. **a,** Associations of proband, paternal and maternal PGS_EA_, PGS_CP_ and PGS_NON_COG_ with quantitative ASD phenotypes in affected offspring in the population model. Effect estimates were obtained from linear regression models including a single polygenic score and meta-analyzed across the discovery and replication cohorts. Color indicates effect size and point size indicates −*log*_10_(*P_adj_*). **b,** Associations of proband, paternal and maternal PGS_EA_, PGS_CP_ and PGS_NON_COG_ with ASD comorbidities in affected offspring in the population model. Points indicate odds ratios and error bars indicate 95% confidence intervals. Asterisks denote Bonferroni-corrected significance levels: *P_adj_* < 0.05 (*); *P_adj_* < 0.01 (**); *P_adj_* < 0.001 (***).

**Figure S3.**
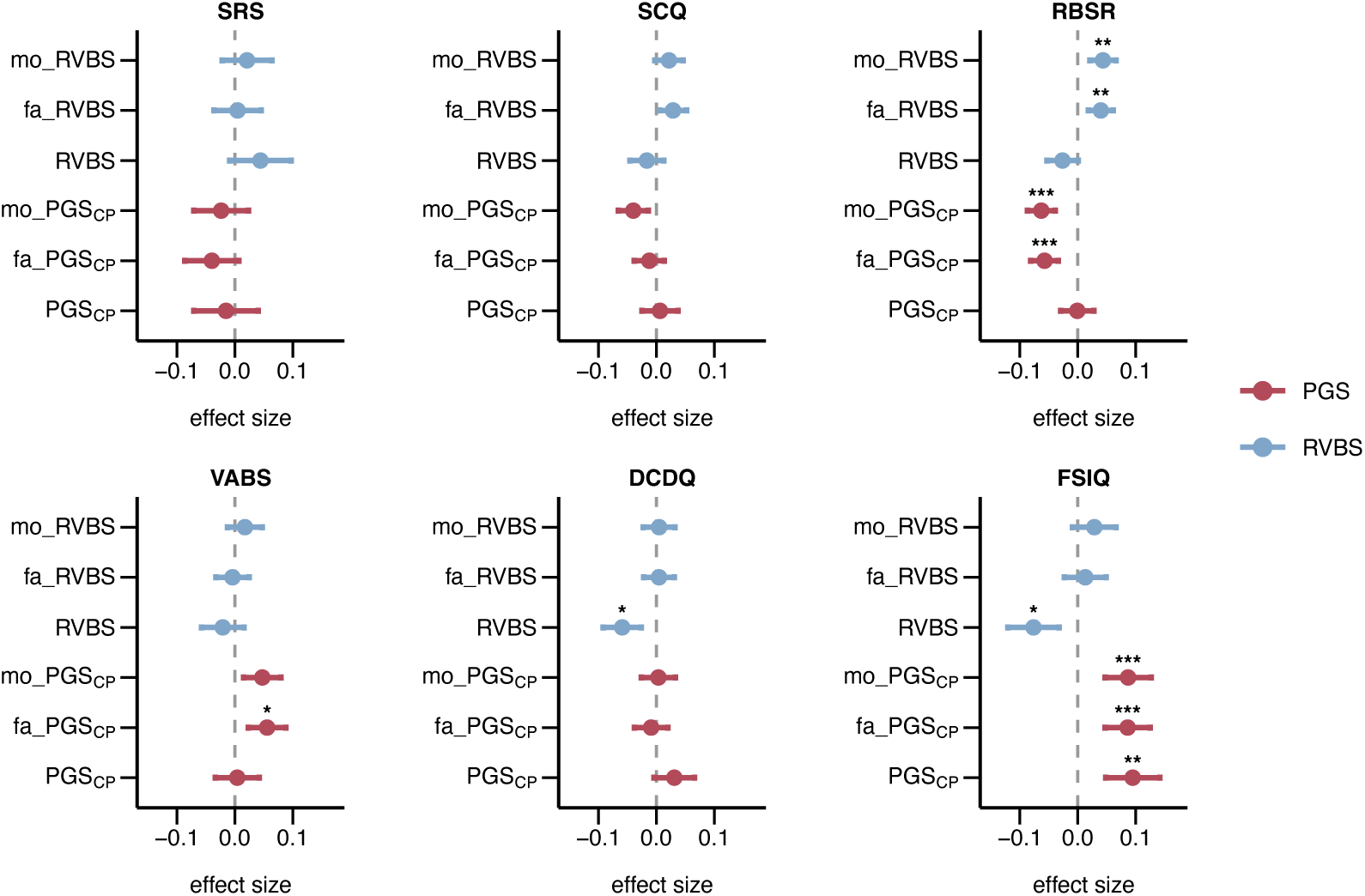
The effects of PGS_CP_ in trio model after adjustment for rare variant burden (RVBS). Effect estimates were obtained from linear regression models and meta-analyzed across the discovery and replication cohorts. Points indicate effect estimates and error bars indicate 95% confidence intervals. Results are shown for all probands, PTV carriers, and PTV non-carriers. Points indicate effect estimates and error bars indicate 95% confidence intervals. Asterisks indicate Bonferroni-corrected significance thresholds: *P_adj_* < 0.05 (*); *P_adj_* < 0.01 (**); *P_adj_* < 0.001 (***).

**Figure S4.**
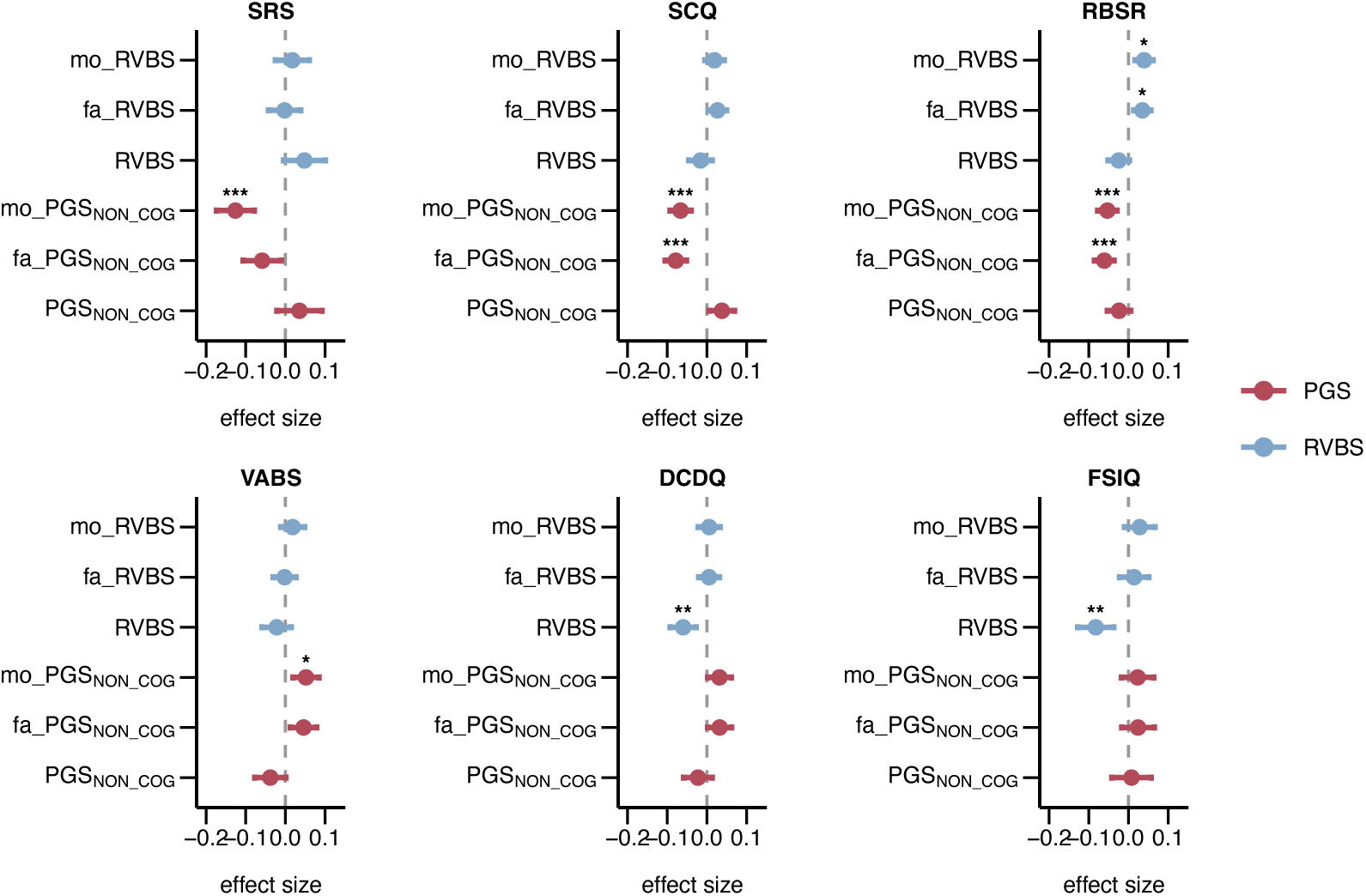
The effects of PGS_NON_COG_ in trio model after adjustment for rare variant burden (RVBS). Effect estimates were obtained from linear regression models and meta-analyzed across the discovery and replication cohorts. Points indicate effect estimates and error bars indicate 95% confidence intervals. Results are shown for all probands, PTV carriers, and PTV non-carriers. Points indicate effect estimates and error bars indicate 95% confidence intervals. Asterisks indicate Bonferroni-corrected significance thresholds: *P_adj_* < 0.05 (*); *P_adj_* < 0.01 (**); *P_adj_* < 0.001 (***).

**Figure S5.**
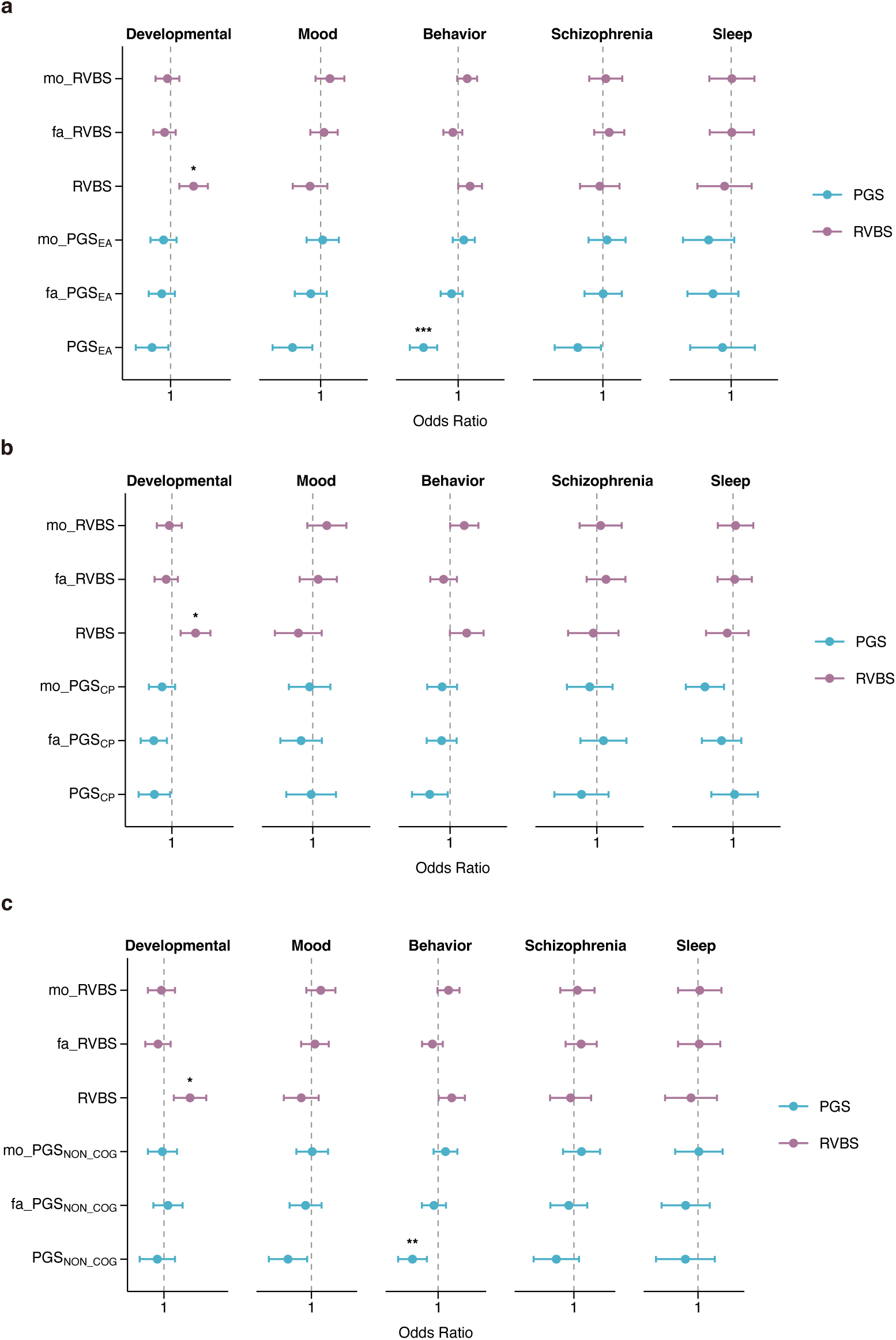
Associations of PGS with ASD comorbidities after adjustment for rare variant burden (RVBS) in the trio model. Associations of PGS (**a,** PGS_EA_, **b,** PGS_CP_, **c,** PGS_NON_COG_) with developmental disorders, mood disorders, behavioral disorders, schizophrenia and sleep disorders were estimated in trio models. Results were meta-analyzed across the discovery and replication cohorts. Points indicate odds ratios and error bars indicate 95% confidence intervals. Asterisks indicate Bonferroni-corrected significance thresholds: *P_adj_* < 0.05 (*); *P_adj_* < 0.01 (**); *P_adj_* < 0.001 (***).

**Figure S6.**
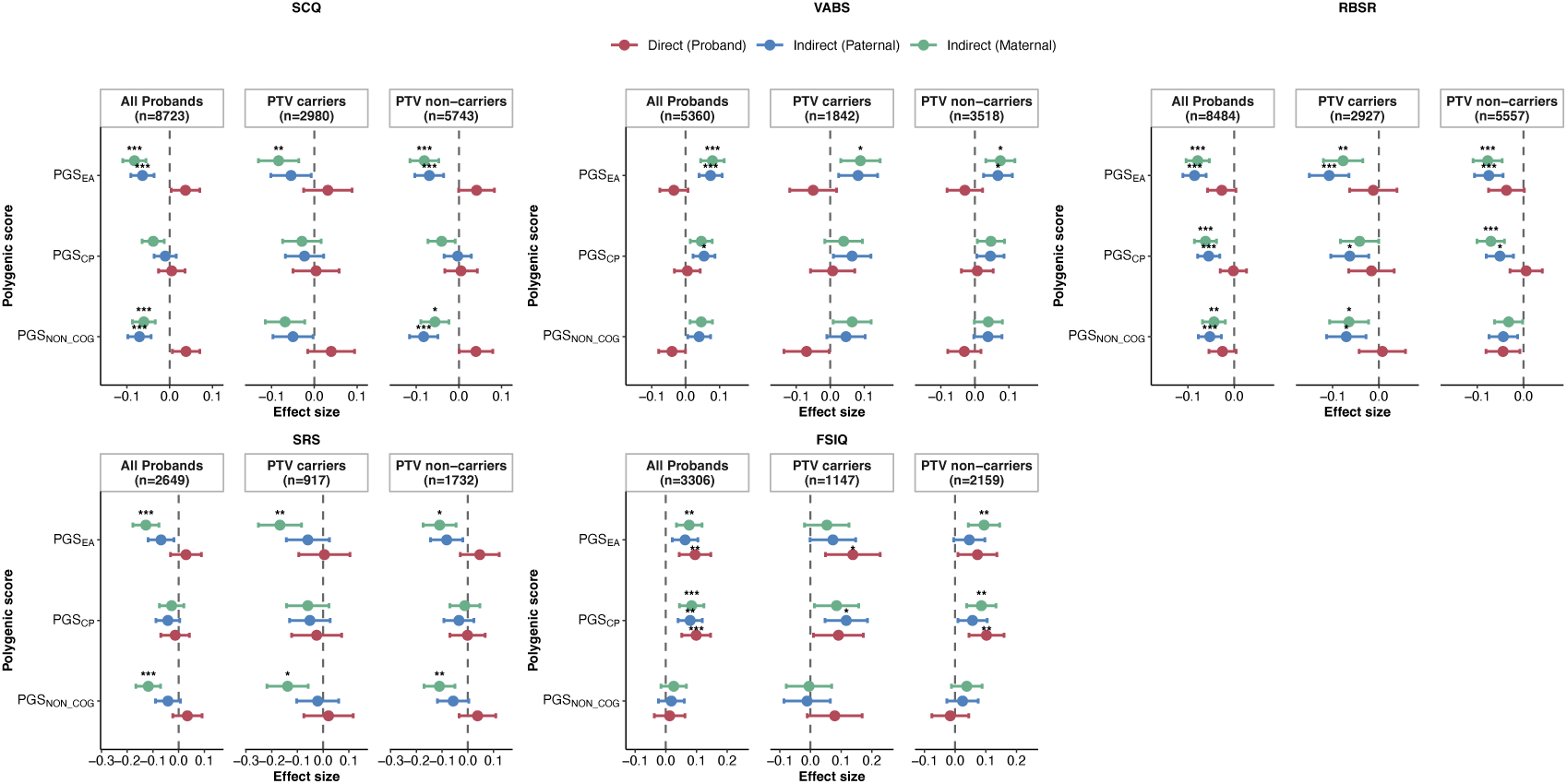
Associations of PGS with ASD phenotypes in the trio model, stratified by SFARI gene PTV carrier status. Associations of PGS_EA_, PGS_CP_ and PGS_NON_COG_ with SCQ, SRS, RBSR, VABS and FSIQ are shown for all probands, PTV carriers, and PTV non-carriers. Points indicate effect estimates and error bars indicate 95% confidence intervals. Asterisks indicate Bonferroni-corrected significance thresholds: *P_adj_* < 0.05 (*); *P_adj_* < 0.01 (**); *P_adj_* < 0.001 (***).

**Figure S7.**
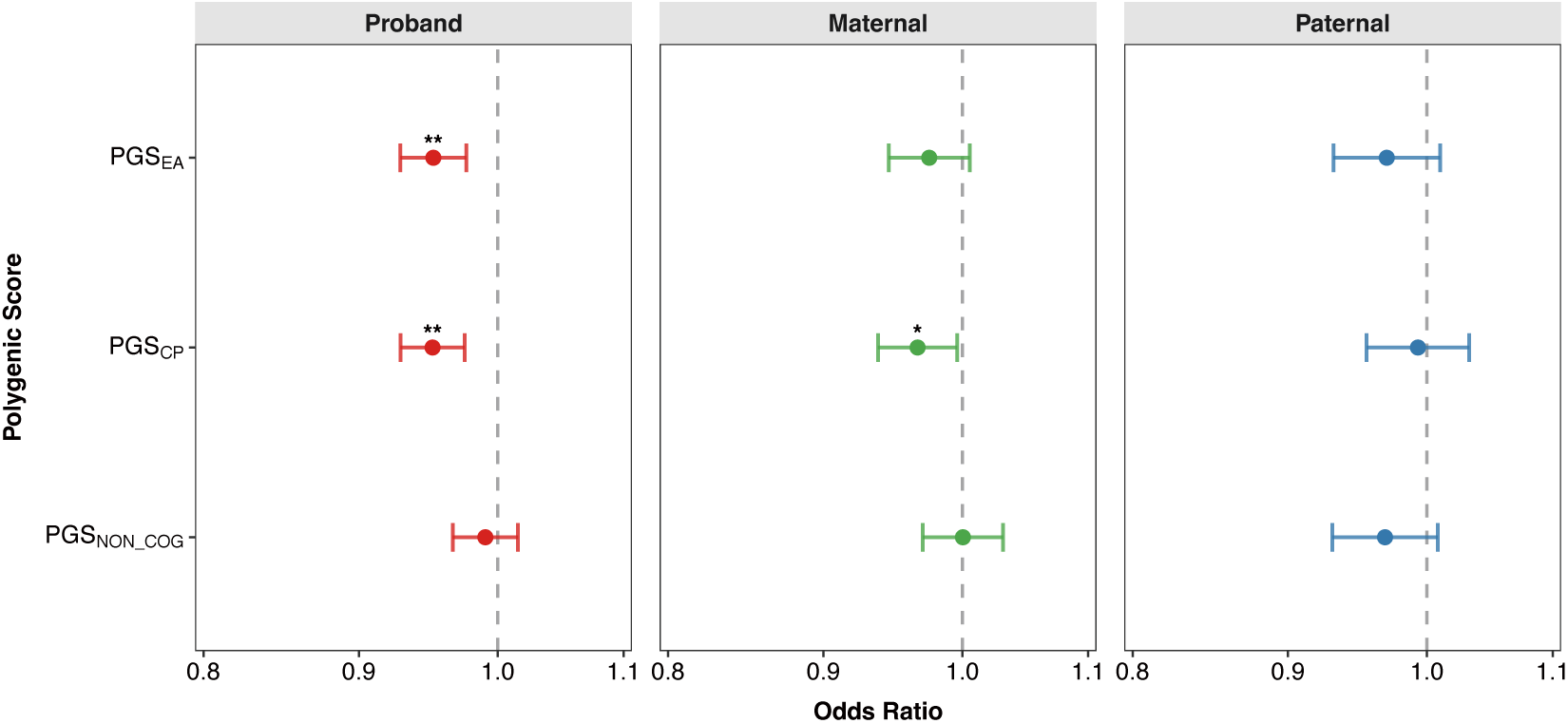
Associations of PGS with prenatal factors in ASD probands. Associations of proband, maternal and paternal PGS_EA_, PGS_CP_ and PGS_NON_COG_ with prenatal factors in ASD probands. Points indicate odds ratios and error bars indicate 95% confidence intervals. Asterisks indicate significance after multiple-testing correction: : *P_adj_* < 0.05 (*); *P_adj_* < 0.01 (**); *P_adj_* < 0.001 (***).

**Figure S8.**
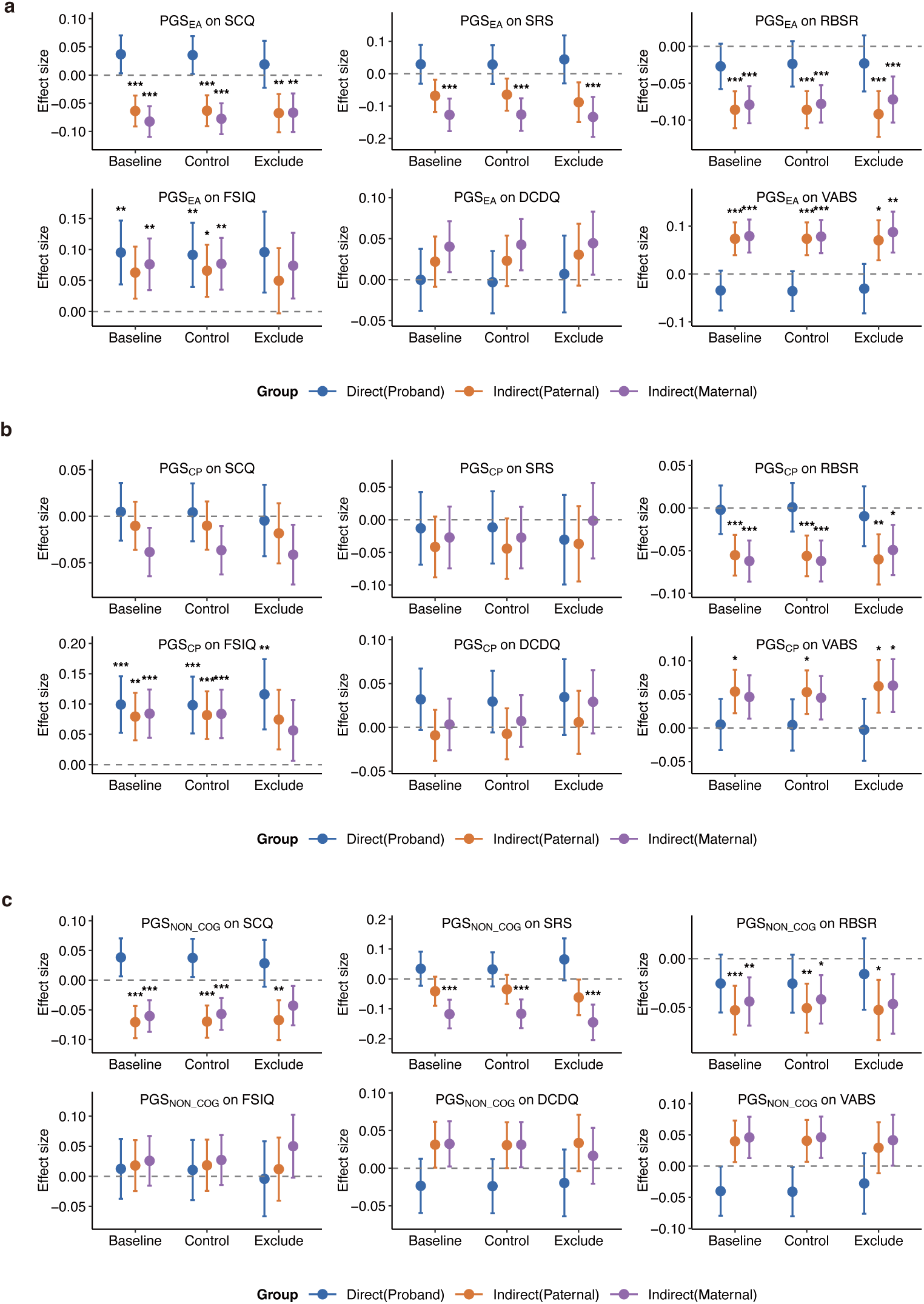
Associations of PGS with ASD phenotypes after accounting for prenatal factors. Associations of PGS (**a,** PGS_EA_, **b,** PGS_CP_, **c,** PGS_NON_COG_) with SCQ, SRS, RBSR, FSIQ, DCDQ and VABS are shown under three analytic strategies: the baseline trio model, additional adjustment for prenatal factors, and exclusion of individuals with prenatal factors. Effect estimates were obtained from trio models including child, paternal and maternal polygenic scores and meta-analyzed across the discovery and replication cohorts. Points indicate effect estimates and error bars indicate 95% confidence intervals. Asterisks indicate significance after multiple-testing correction: *P_adj_* < 0.05 (*); *P_adj_* < 0.01 (**); *P_adj_* < 0.001 (***).

**Figure S9.**
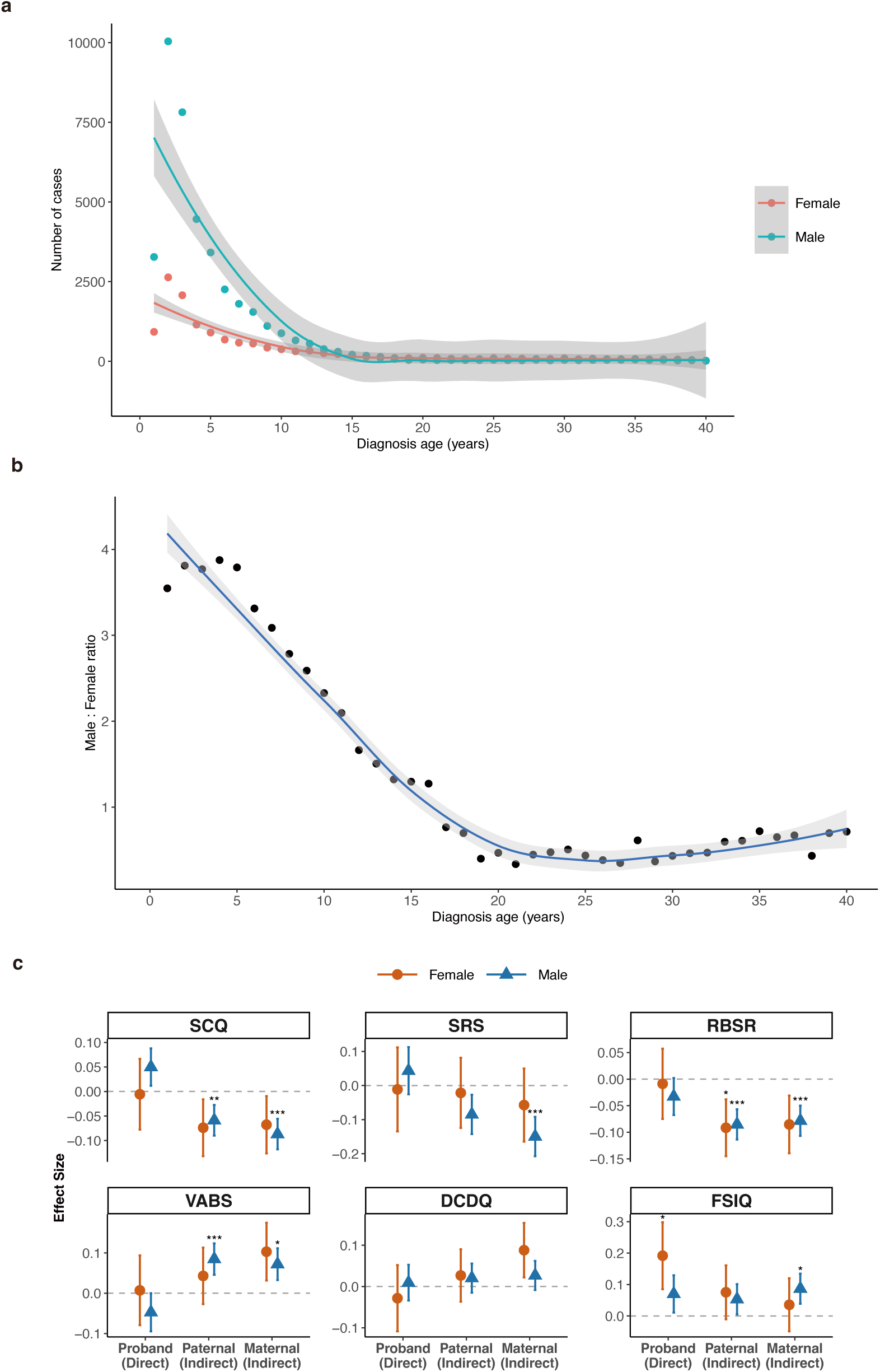
Sex distribution by age at diagnosis and sex-stratified trio-model associations of educational attainment-related polygenic scores with ASD phenotypes. **a,** Number of male and female ASD cases across age at diagnosis. Points indicate observed counts and curves indicate loess-smoothed trends with shaded 95% confidence intervals. **b,** Male:female ratio across age at diagnosis. Points indicate observed ratios and the curve indicates the loess-smoothed trend with shaded 95% confidence intervals. **c,** Associations of PGS_EA_ with SCQ, SRS, RBSR, VABS, DCDQ and FSIQ in sex-stratified trio models. Results are shown separately for males and females. Points indicate effect estimates and error bars indicate 95% confidence intervals. Asterisks indicate significance after multiple-testing correction: *P_adj_* < 0.05 (*); *P_adj_* < 0.01 (**); *P_adj_* < 0.001 (***).

## Notes

### Competing Interest Statement

The authors have declared no competing interest.

### Author Declarations

The genomic and phenotypic data for the SPARK study (https://www.sfari.org/resource/spark/) are available at SFARI Base (https://base.sfari.org). The study was reviewed with ethical approval given by the Biomedical Ethics Committee of Peking University (No. PUIRB-YS2023128).

